# A Modeling Analysis of Strategies for Allocating Emergency Department HIV and Syphilis Screening Funds in California

**DOI:** 10.64898/2026.09.17.26363340

**Authors:** Indrani Guzman Das, Ribhav Gupta, Christopher L. Bennett

## Abstract

**Introduction:** Using an open enrollment process in 2023, California funded 28 emergency departments (EDs) to implement or expand HIV and syphilis screening programs. California’s 2026 state budget now commits an additional $60 million, over six years, to renew support for this screening. How renewed funds are allocated will determine which communities they reach. This analysis establishes a framework derived from public information to simulate and compare several funding strategies, highlighting the tradeoffs across disease burden, geographic reach, and social vulnerability.

**Methods:** Publicly available data were used to construct ED-level profiles for the 282 EDs in the 35 California counties with populations exceeding 100,000. Profiles were then linked to corresponding HIV and syphilis burden (z-score), measures of social vulnerability for the surrounding service area , and socioeconomic and demographic characteristics of the population each ED serves. At a 28 ED award scale, allocation strategies (random selection, prior 2023 awardees, federal priority-county targeting, ranking on burden alone, and a composite of burden, vulnerability, and visit volume) were then compared, simulating ED participation across 100,000 Monte Carlo draws at 70% participation. Sensitivity analyses included varied (50% and 90%) participation and a larger (n=64) ED award scale. Analyses were conducted in 2026.

**Results:** No single strategy performed best. Ranking on burden alone reached the highest mean county burden but only 4 of 35 counties, whereas a composite approach reached 8 counties, the highest social vulnerability, and the largest share of patients who identified as Hispanic. Every targeted strategy performed better than both random selection and the prior 2023 awardees on burden but reached fewer counties than either. In sensitivity analyses, the tradeoffs between burden and reach persisted across participation rates and at the 64-ED scale.

## INTRODUCTION

Emergency departments (EDs) are a primary point of contact with the healthcare system for a growing share of the US population, delivering close to half of all hospital-associated medical care.^1^ EDs are also tasked with providing a growing amount of primary and preventive care for Americans who are either uninsured or face barriers to alternative forms of care.^2^ Despite the role that EDs play in healthcare access, particularly among communities that disproportionately face social and structural determinants of health associated with poorer health outcomes, they remain underused in national efforts to screen for sexually transmitted infections (STIs). The need is substantial; HIV remains a persistent challenge in the US. Roughly one in seven Americans living with HIV remain unaware of their infection.^3,4^ In parallel, syphilis has resurged over the past decade, forming a distinct syndemic with HIV.^5^

Given these growing public health challenges, in 2019 the Centers for Disease Control and Prevention (CDC) launched the *Ending the HIV Epidemic in the U.S*. (EHE) Initiative, prioritizing high-burden jurisdictions with the goal of reducing new infections through early diagnosis.^6^ EDs in these jurisdictions disproportionately serve the vulnerable populations EHE emphasizes, persons who are uninsured or living in poverty and communities disproportionately impacted by multiple STI epidemics.^7^ Additionally, we previously reported that the subset of EDs in priority jurisdictions affiliated with teaching hospitals may be particularly well positioned to contribute to EHE’s goals given both their unique social mission and patient populations along with their advanced clinical capabilities.^7,8^

California carries one of the nation’s highest HIV burdens and, in 2023 through the California Department of Public Health, launched the ED Syphilis/HIV/hepatitis C (HCV) Screening Program (EDSP). Using an open application process, rather than an explicit targeting framework, EDSP allocated $13.1 million to 28 California EDs.^9^ California’s recently finalized state budget includes an additional $60 million commitment over the next six years to further support ED-based HIV, hepatitis C, and syphilis screening and linkage to care programs.^10^ This renewed funding represents a critical opportunity for California to build on its initial investment into local EDs and refine how resources might be most effectively allocated. However, to our knowledge, no study has evaluated how alternative allocation criteria might change the communities in California these ED-based funds could reach. As such, the objectives of this policy analysis were to simulate and compare potential strategies for allocating renewed funding across California EDs, to evaluate the tradeoffs each would produce in reaching different communities, and to offer a framework derived solely on publicly available information that could be adapted and applied for future efforts, for other efforts and for different communities.

## METHODS

### Data Sources

We constructed facility-level profiles for the 282 general acute-care EDs in California with more than 500 annual visits using publicly available data from California’s Department of Health Care Access and Information.^11^ Profiles included annual visit volume, teaching hospital status, county location, and Medical Service Study Area (MSSA). In line with updated guidance on the reporting of race and ethnicity, profiles also included socioeconomic and demographic characteristics of patients served by each ED; MSSA represents a sub-county geographic unit, in part, used to identify medically underserved populations.^12^ We then restricted the sample of EDs to those in the 35 California counties with populations exceeding 100,000. These California counties contained 91% of qualifying EDs and 97% of the state’s population. Smaller counties, disproportionately rural and frontier, were excluded because their annual case counts produced unstable per-capita rates. EDs were then linked to the annual number of HIV diagnoses and primary and secondary syphilis cases reported in its county using CDC’s AtlasPlus surveillance system. To our knowledge, county-level estimates are the smallest geographic level at which diagnoses are publicly reported.^13^ Most recent, 2025, data for HIV were reported as preliminary, prompting use of 2024 data; 2023 data for syphilis represented the most recent data available.

We then obtained census-tract level information on overall social vulnerability from CDC’s Social Vulnerability Index (SVI). Census tracts along with corresponding SVI scores and estimated total population, via Federal Information Processing Standards Codes, were then assigned to their corresponding MSSAs. To create an MSSA-level population weighted average of social vulnerability, each tract within an individual MSSA was identified, and its SVI score was multiplied by its total population. These weighted values were summed across all tracts in the MSSA, dividing by the MSSA’s total population. Tracts with suppressed SVI data were excluded. Each ED was then assigned the population-weighted SVI score of the MSSA in which it was located.

Finally, individual EDs that received funding under the original 2023 allocation were identified through the EDSP’s published list of awardees. Twenty-seven of the 28 were matched successfully. One awardee was not present in the available data and one awardee was in a county below the population threshold; the final set of prior awardees included 26 EDs. The Stanford University Institutional Review Board deemed this study exempt given that it represented a secondary analysis of publicly available, aggregate, data.

### Syndemic Burden and Vulnerability Measures

County-level syndemic burden was defined as the average of two standardized (z-scored) rates (county-level HIV diagnosis rate and county-level primary and secondary syphilis rates, both per 100,000 population), each standardized across all California counties with an available rate for that measure. Of the 35 counties included in our analysis, only one (El Dorado) had suppressed HIV rates. For this county, syndemic burden score reflected the syphilis rate alone rather than an average of both, **Supplemental Figure 1**. Sensitivity analyses incorporating an imputed multi-year rolling average of HIV for this county did not materially change outcomes. Finally, five hypothetical allocation strategies were simulated.

### Funding Allocation Strategies

Strategy A represented a process where eligible EDs were drawn at random, without replacement, across 100,000 simulated draws. Although likely not a prediction of actual outcomes given the expectation that applicants would more often originate from institutions with grant-writing capacity, existing screening infrastructure, and other factors a random draw would not capture, it represented a mathematical base case reference point characterizing what no targeting criterion might produce. Because county-level syndemic burden is assigned identically to every ED in its corresponding county, drawing EDs at random over-samples high-burden, ED-dense, counties. As such, random selection reaches a mean burden above zero rather than the zero a county-level draw would produce. We reported this ED-level value as the random benchmark given funding is allocated to EDs, not counties, **Supplemental Figure 2**.

Strategy B represented a selection process grounded in EHE’s geographic framework, further emphasizing the subset of California EDs within priority jurisdictions with a teaching hospital affiliation. EDs were ranked by four sequential criteria: (1) location in an EHE priority jurisdiction (yes before no); (2) affiliation with a teaching hospital (yes before no); (3) county syndemic burden z-score (descending); and (4) annual ED visit volume (descending) to break ties among EDs identical on the first three criteria, **Supplemental Figure 3**.

Strategy C represented the most direct application of disease burden, assigning higher ranking to EDs by county syndemic burden alone without incorporating EHE’s priority-jurisdiction framework. EDs were ranked by three sequential criteria: (1) county syndemic burden z-score (descending); (2) teaching-hospital affiliation (yes before no); and (3) annual ED visit volume (descending) to break ties among EDs identical on the first two criteria.

Strategy D combined three factors into a single score for each ED: county syndemic burden, MSSA-level social vulnerability, and ED visit volume. Each factor was standardized (z-scored) across the full pool and averaged with equal weight. Vulnerability was measured at the MSSA level rather than the county level, since county-level vulnerability, like county-level burden, is identical for every ED sharing a county. EDs were then ranked by this composite score. To assess how sensitive this ranking was to the equal-weighting choice incorporated here, sensitivity analyses testing three alternative weightings, emphasizing syndemic burden, social vulnerability, or visit volume were incorporated under the primary participation rate, **Supplemental Figure 4**.^14^

Strategy E assumed continued funding to the same group of EDs selected in the initial 2023 iteration, limited to the 35-county analytic pool. Given that future funding could include a larger number of EDs, a Strategy F was also included in sensitivity analyses as an additional comparator at a larger award scale. Strategy F assumed that prior awardees would continue to be funded with remaining awards filled with not-yet-funded EDs, selecting new EDs for funding by Strategy D’s composite score.

### Modeling participation uncertainty and Outcomes Reported

Each strategy was evaluated at two award scales: 28 EDs (which holds the number of funded EDs at the 2023 level) in the primary analysis and 64 EDs in the sensitivity analysis (which holds the per-ED funding rate at the 2023 level while expanding the cohort to approximate one $30 million installment). Given the anticipation that not every ranked ED can be expected to apply for and successfully implement funding, we modeled imperfect participation for the ranked strategies (B-D and F), where each ED was assigned a probability of participating if selected, and a funding opportunity passed to the next-ranked ED otherwise. This included conservative (50%), realistic (70%), and optimistic (90%) participation probabilities, incorporating 70% in the primary analyses; Strategy F was modeled under the same three participation probabilities as Strategies B-D. Each participation scenario was simulated across 100,000 draws. For each strategy we measured mean county burden (z-score) reached, number of counties reached (of 35), mean MSSA social vulnerability, along with the number and proportion of ED visits reached overall and among populations emphasized in multiple state and federal HIV initiatives (e.g., persons who were homeless or uninsured, or identified as Black or Hispanic). For each measure, uncertainty was summarized with Monte Carlo simulation intervals.

## RESULTS

Across the measures analyzed, each strategy demonstrated distinct strengths, **Table 1**. California’s 2023 EDSP awardees (Strategy E) reached vulnerable, at-risk, communities with greater disease burden than random selection. Although every targeted strategy (Strategies B through D) reached higher disease burden than random selection (Strategy A), no single strategy performed best. Random selection (Strategy A) reached a mean county syndemic burden of 0.25 (95% simulation interval [SI] 0.01–0.50) across 15 (12–19) of the 35 counties. Among the targeted strategies, ranking EDs directly by county burden (Strategy C) reached the highest mean burden, 1.24 (1.10–1.36), but only 4 (4–4) counties. The priority-county plus teaching strategy (Strategy B) reached a mean burden of 0.89 (0.73–1.05) across 7 (6–8) counties. The equal-weighted composite score (Strategy D) reached a mean burden of 0.85 (0.75–0.93) across 8 counties (6–10). California’s 2023 awardees (Strategy E) reached a mean burden of 0.46 across 13 counties, above random selection but below every other simulated targeted strategy. Random selection (15 counties) and the 2023 awardees (13 counties) reached more counties than any targeted strategy (4 to 8 counties).

**Table 1.** Performance of simulated strategies against actual 2023 awardees.

| Measure | A. Simple random benchmark | B. Priority county + teaching | C. Direct burden rank | D. Composite | E. Actual 2023 awardees |
| --- | --- | --- | --- | --- | --- |
| Counties reached (of 35) | 15 (12-19) | 7 (6-8) | 4 (4-4) | 8 (6-10) | 13 |
| Mean county burden (z-score) | 0.25 (0.01-0.50) | 0.89 (0.73-1.05) | 1.24 (1.10-1.36) | 0.85 (0.75-0.93) | 0.46 |
| Mean social vulnerability (MSSA SVI, 0-1) | 0.51 (0.43-0.58) | 0.51 (0.46-0.55) | 0.54 (0.50-0.59) | 0.74 (0.70-0.77) | 0.68 |
| Homeless-patient visits reached, n (%) | 43,508 (2.9%)<br>[1.9-4.3%] | 109,870 (4.8%)<br>[3.6-5.8%] | 76,904 (4.4%)<br>[3.2-5.5%] | 86,638 (3.9%)<br>[3.0-4.7%] | 83,351 (5.2%) |
| Uninsured visits reached, n (%) | 72,318 (4.9%)<br>[3.9-5.9%] | 117,976 (5.1%)<br>[4.5-5.7%] | 86,242 (4.9%)<br>[4.4-5.5%] | 114,141 (5.1%)<br>[4.5-5.8%] | 96,289 (6.0%) |
| Black-patient visits reached, n (%) | 149,798 (10.0%)<br>[6.8-13.9%] | 345,761 (15.0%)<br>[12.8-16.9%] | 235,347 (13.4%)<br>[11.0-15.6%] | 262,314 (11.8%)<br>[10.1-13.4%] | 239,403 (15.0%) |
| Hispanic-patient visits reached, n (%) | 663,808 (44.5%)<br>[36.9-52.2%] | 1,058,137 (45.8%)<br>[41.5-50.4%] | 897,434 (51.1%)<br>[47.0-55.2%] | 1,276,788 (57.3%)<br>[53.4-60.7%] | 815,687 (51.1%) |
| ED visit volume reached, n (%) | 1,489,500 (9.9%)<br>[7.8-12.1%] | 2,309,651 (15.4%)<br>[13.9-17.0%] | 1,753,024 (11.7%)<br>[10.0-13.4%] | 2,228,592 (14.9%)<br>[13.7-16.0%] | 1,595,707 (10.6%) |

**Legend:** Strategies A-E at the 28-emergency department funding scale. Ranked Strategies B-D are modeled at 70% participation. Values are presented as means with 95% simulation interval [SI] across 100,000 draws.
**Abbreviations:** ED (emergency department), MSSA (Medical Service Study Area), SVI (social vulnerability index)

Differences in geographic reach also tracked with how burden was measured. Although incorporating teaching hospital affiliation and ED visit volume, Strategy C centered on awarding by county-level burden; expectedly this approach filled available awards with EDs from a small number of high-burden counties. This contrasted with the composite score approach (Strategy D) that incorporated social vulnerability and ED visit volume, distinguishing EDs within counties; this approach reached the highest mean social vulnerability (0.74 [0.70–0.77]), followed by the 2023 awardees (0.68); Strategies B and C (0.51 and 0.54, respectively) reached communities no more vulnerable than those reached by random selection (0.51). Similarly, characteristics of the population reach also differed by strategy. Strategy D reached the largest share of visits by persons who identify as Hispanic (57.3% [53.4–60.7%]) and Strategy B reached the largest volume of ED visits overall (approximately 2.3 million visits, or 15% of statewide volume).

Although 2023 awardees reached the largest shares of homeless (5.2%) and uninsured (6.0%) visits, the volume of ED visits reached was similar to that of random selection.

These findings persisted under several different assumptions of imperfect participation (**Figure 1** and **Supplemental Table 1**). At every modeled participation rate, Strategy C reached the highest mean burden and every targeted strategy exceeded both random selection and the 2023 awardees (e.g., mean burden under Strategies B through D ranged from 0.82 to 1.13 at 50% participation and from 0.78 to 1.34 at 90% participation). Findings from sensitivity analyses assuming a larger, 64-ED, award scale were similar (**Supplemental Table 2** and **Supplemental Figure 5**). At this scale, Strategy F, which continued funding for the 2023 awardees and filled the remaining awards with new EDs, reached a mean burden of 0.71 (0.63–0.78) across 16 counties (14–19), above the 2023 awardees on burden and matching Strategy D on reach.

**Figure 1.**
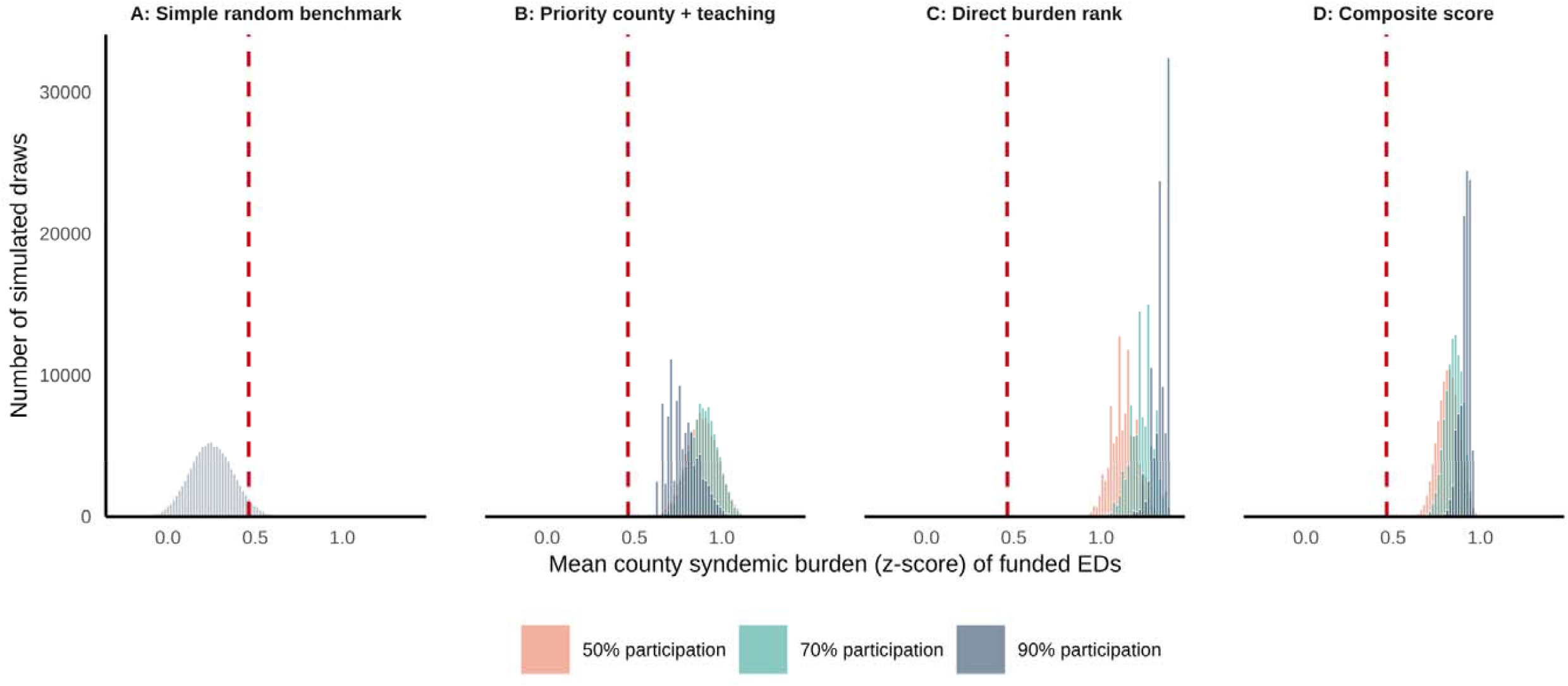
Mean county burden by strategy and participation rate, compared to random selection and the 2023 awardees. **Legend**: Distributions of mean county syndemic burden for Strategies B–D at 50%, 70%, and 90% participation, 100,000 draws each. Presented at the 28-emergency department award scale. Strategy A is represented by a gray distribution of 100,000 random draws. Strategy E is represented in each pane by a red, dashed, vertical reference line. **Abbreviations**: ED (emergency department)

The absence of a dominant strategy also held across different approaches to weighting Strategy D’s composite score (**Figure 2**). Burden-first weighting reached the highest mean burden (1.20) across the fewest counties (5) while equity-first weighting reached the most counties (10) at a mean burden of 0.76. Equal (0.85 across 8 counties) and efficiency-first (0.67 across 9 counties) weightings fell in between on reach. No weighting reached both a higher mean burden and more counties than the equal-weighted composite. In sensitivity analyses, assuming a 64-ED award scale, findings were similar (**Supplemental Figure 6**).

**Figure 2.**
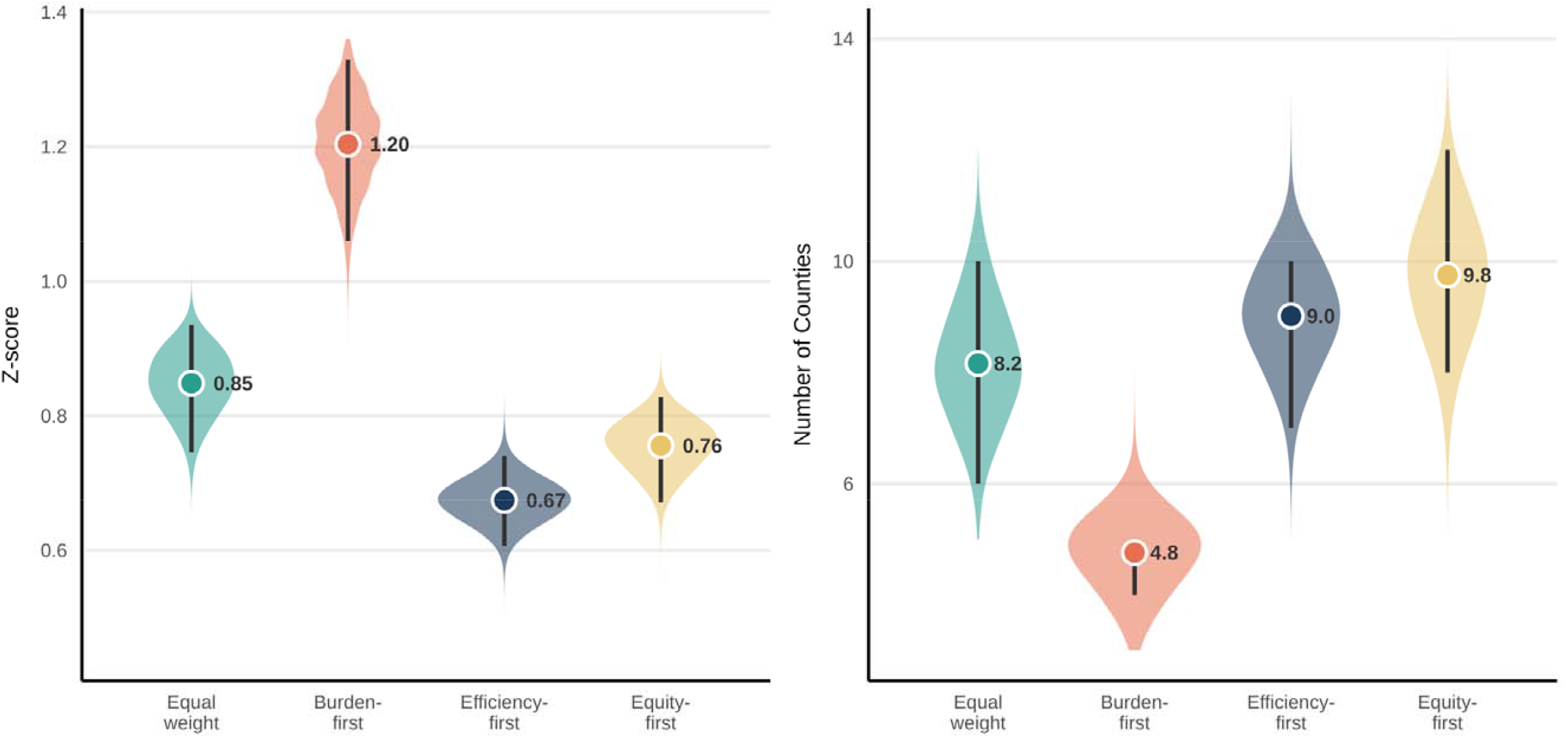
Impact of re-weighting the composite score. **Legend**: Mean county burden (left; z-score) and counties reached (right; of 35) under four weightings of Strategy D’s composite at 70% participation. Violins show the full distribution across 100,000 draws. The point marks the mean with corresponding vertical bars representing the 95% simulation interval. Composite weightings of burden, vulnerability, and volume: equal (1/3, 1/3, 1/3), burden-first (0.6, 0.2, 0.2), equity-first (0.2, 0.6, 0.2), and efficiency-first (0.2, 0.2, 0.6). **Abbreviations**: ED (emergency department)

## DISCUSSION

In this study we present a framework assessing the outcomes of multiple potential funding strategies using exclusively public data. We identified cohorts of EDs that might receive renewed state-level, ED-based, funding for HIV and syphilis screening. Under each strategy we compared the disease burden, geographic reach, and social vulnerability of the communities each would reach. To our knowledge, this is the first analysis simulating alternative strategies for allocating California’s ED-based screening funds, quantifying the tradeoffs among them. Our analysis has three core findings. First, developing a framework to assess funding strategies using exclusively public data was feasible. The strategies considered reached higher mean burden than both random allocation and California’s prior 2023 open-application awardee process. This gain came at a cost in geographic reach, as every targeted strategy funded EDs in fewer counties than either. Second, no single strategy performed best on disease burden, geographic reach, and social vulnerability at the same time. Third, because these tradeoffs persisted across weightings, award scales, and participation assumptions, the choice among strategies is a decision about which objective to prioritize.

The way these funds are awarded itself is a policy decision. An open application does not avoid a choice among priorities; it makes that choice implicitly through which EDs apply. In this analysis we found that re-funding the 2023 awardees corresponded to a high-vulnerability, geographically broad set of EDs, but one that reached lower disease burden than every targeted strategy at the same 28-ED scale. Specifying objectives, whether targeting disease burden, reach, social vulnerability or a defined combination, may make the allocation explicit and allow it to be evaluated against that objective.

These considerations are not unique to California. Jurisdictions across the US are tasked with allocating limited screening funds (e.g., EHE, the Ryan White program, state or federal viral hepatitis efforts, or other coordinated responses) for other transmissible infectious diseases; these jurisdictions could apply similar frameworks to aid in award allocation. Our findings here also indicate a limitation of relying on federal priority-county designations as the sole targeting criterion; EHE’s framework only partially aligned with county burden and social vulnerability at a sub-national level. This finding is consistent with prior, national, work from our group.^15^ Strategies built exclusively on this designation might inefficiently reach counties at a state-level. This in turn could omit higher burden EDs outside priority counties. With public health budgets under increasing strain, namely for HIV-related efforts, every screening dollar counts. An explicit framework lets funders direct each dollar toward a stated objective.

### Limitations

This work has several limitations and should be interpreted accordingly. First, the 2023 program and 2026 renewal center on initiatives targeting the syndemic of HIV, HCV, and syphilis. The definition of syndemic incorporated here is narrower and limited by the absence of data on HCV. Further, HIV and syphilis cases are reported at the county level. Although estimating and modeling sub-county level burden of both are the focus of ongoing work, more granular ED-or census tract-level estimates are not available. Given prior work demonstrates a correlation between social vulnerability and HIV risk, we approximated sub-county burden through MSSA-level social vulnerability.^15^ Second, the ability to implement screening likely varies across EDs. Boarding, crowding, and other operational priorities will likely limit both real-world participation and implementation. The teaching and high-volume EDs best positioned for funding are also likely the most boarding-and crowding-constrained operationally.^16^ As such, we modeled uncertainty using several probabilities of participation. Third, the outcomes evaluated here represent potential reach rather than the clinical outcomes each strategy might ultimately achieve. Modeling differences across EDs with regards to screening eligibility, uptake, test positivity, or costs were beyond the scope of this analysis. As such, these findings should be interpreted as comparisons of allocation strategies and their associated trade-offs, rather than as projections of the number of infections detected, patients linked to care, or health outcomes achieved.^17^ Fourth, the allocation scenarios considered here assume a fixed number of awards (either 28 or 64 EDs funded) and an equal level of funding per ED. The current renewal program may ultimately fund a different number of EDs or vary award amounts according to facility size, existing infrastructure, or implementation needs. Accordingly, this analysis should be interpreted as practical framework derived from publicly available information by which policymakers could compare different facility-selection strategies under a fixed award structure, rather than as a comprehensive optimization of how funding should be distributed. Fifth, the findings presented here incorporate assumptions in both definitions and weighting choices.

Although sensitivity analyses were used to examine several key assumptions, other reasonable specifications could produce different rankings.

## CONCLUSION

ED screening funds can be directed toward higher disease burden, broader geographic reach, or greater social vulnerability but not all three simultaneously. Built entirely on public data, the framework and approach demonstrated here for California could be useful in informing other jurisdictions tasked with allocating similar public health funding.

## Data Availability

All data are already publicly available

https://data.chhs.ca.gov/dataset/hospital-emergency-department-characteristics-by-facility-pivot-profile

https://www.atsdr.cdc.gov/place-health/php/svi/svi-data-documentation-download.html

https://www.cdc.gov/nchhstp/about/atlasplus.html

## Notes

**Financial Support:** CLB reports support from the National Institute of Allergy and Infectious Diseases (L30AI178800 and K08AI181642).

**Conflicts of Interest:** None declared

### Competing Interest Statement

The authors have declared no competing interest.

### Author Declarations

This study used only openly available aggregate, de-identified, data.

