## Supplemental Figures for "A Modeling Analysis of Strategies for Allocating Emergency Department HIV and Syphilis Screening Funds in California"

**Supplemental Figure 1.** Distribution of county-level syndemic burden z-scores

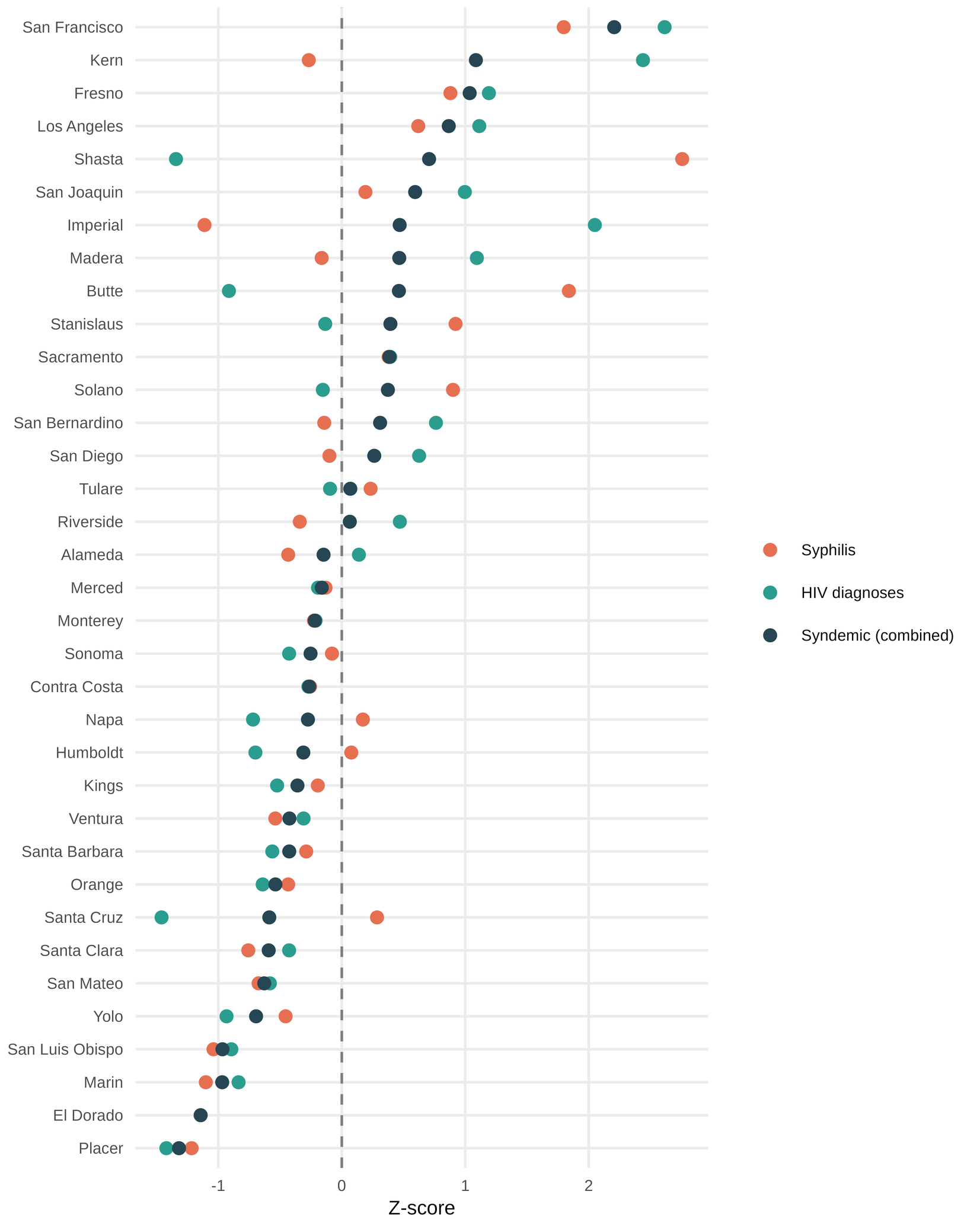

**Legend**: County-level syndemic burden (i.e., the mean of standardized HIV and primary/secondary syphilis rates) for the restricted pool of 35 counties, ranked. El Dorado County (second from bottom) had a suppressed HIV rate; its burden reflects syphilis alone. A multi-year rolling-average HIV imputation did not change results, and its two emergency departments ranked 266–279 of 282, below any funding line.

**Supplemental Figure 2.** Expected burden under emergency department-level versus county-level random sampling

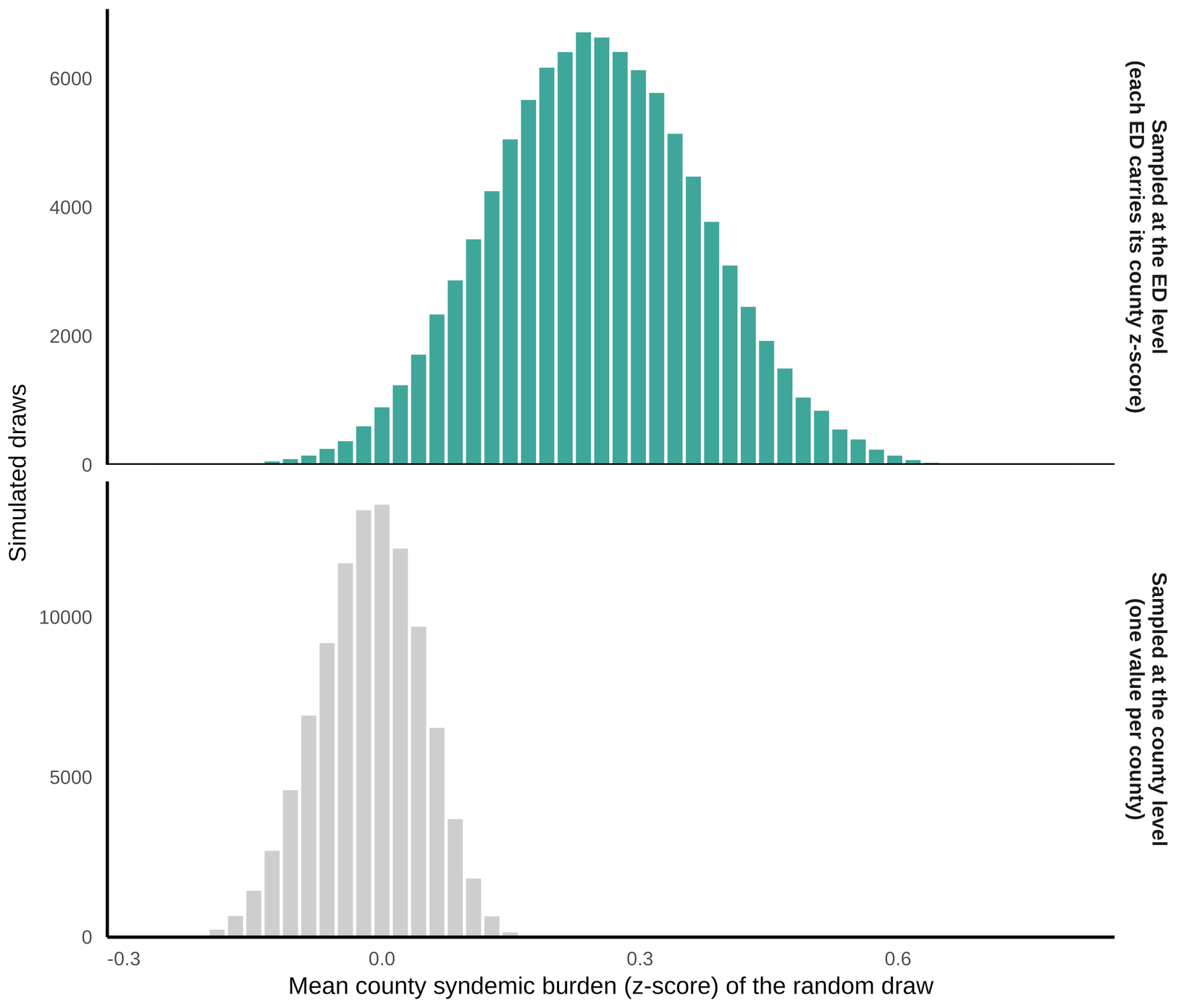

**Legend**: 100,000 random draws of 28. Sampling at the emergency department level yields a mean county-burden z-score modestly above zero (~0.25) given that higher-burden counties contain more emergency departments. Sampling at the county level centers near 0. Because funding is allocated to emergency departments, the emergency department-level value is the reported Strategy A benchmark.

**Abbreviations**: ED (emergency department)

**Supplemental Figure 3.** Strategy B ranking stepwise application of the four ordering criteria

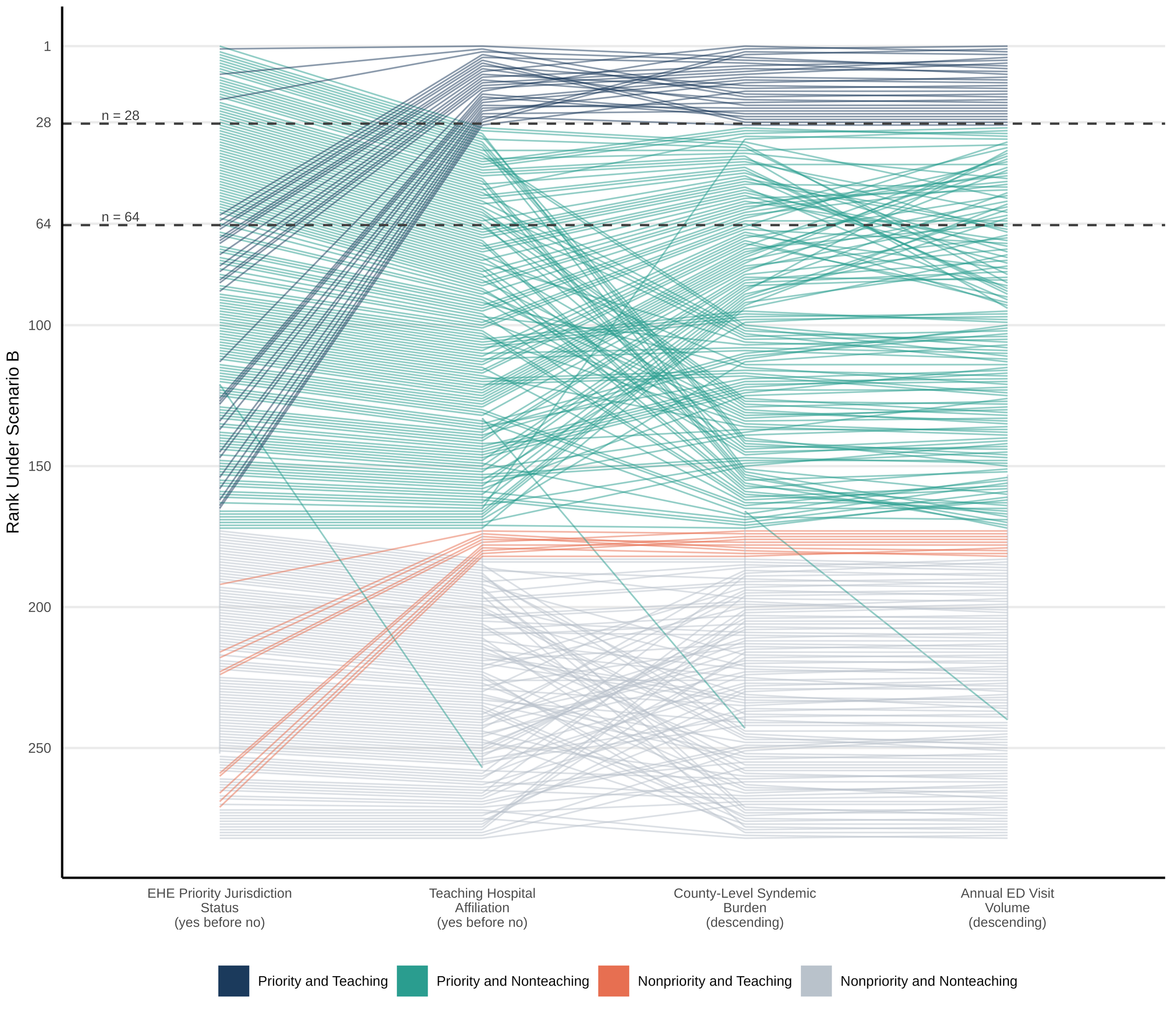

**Legend**: All 282 emergency departments included, re-ranked as each criterion is applied. Horizontal lines mark the n=28 and n=64 funding award scale cutoffs.

**Abbreviations**: ED (emergency department), EHE (Ending the HIV Epidemic in the US Initiative)

**Supplemental Figure 4.** Strategy D composite scores under four different approaches to weighting
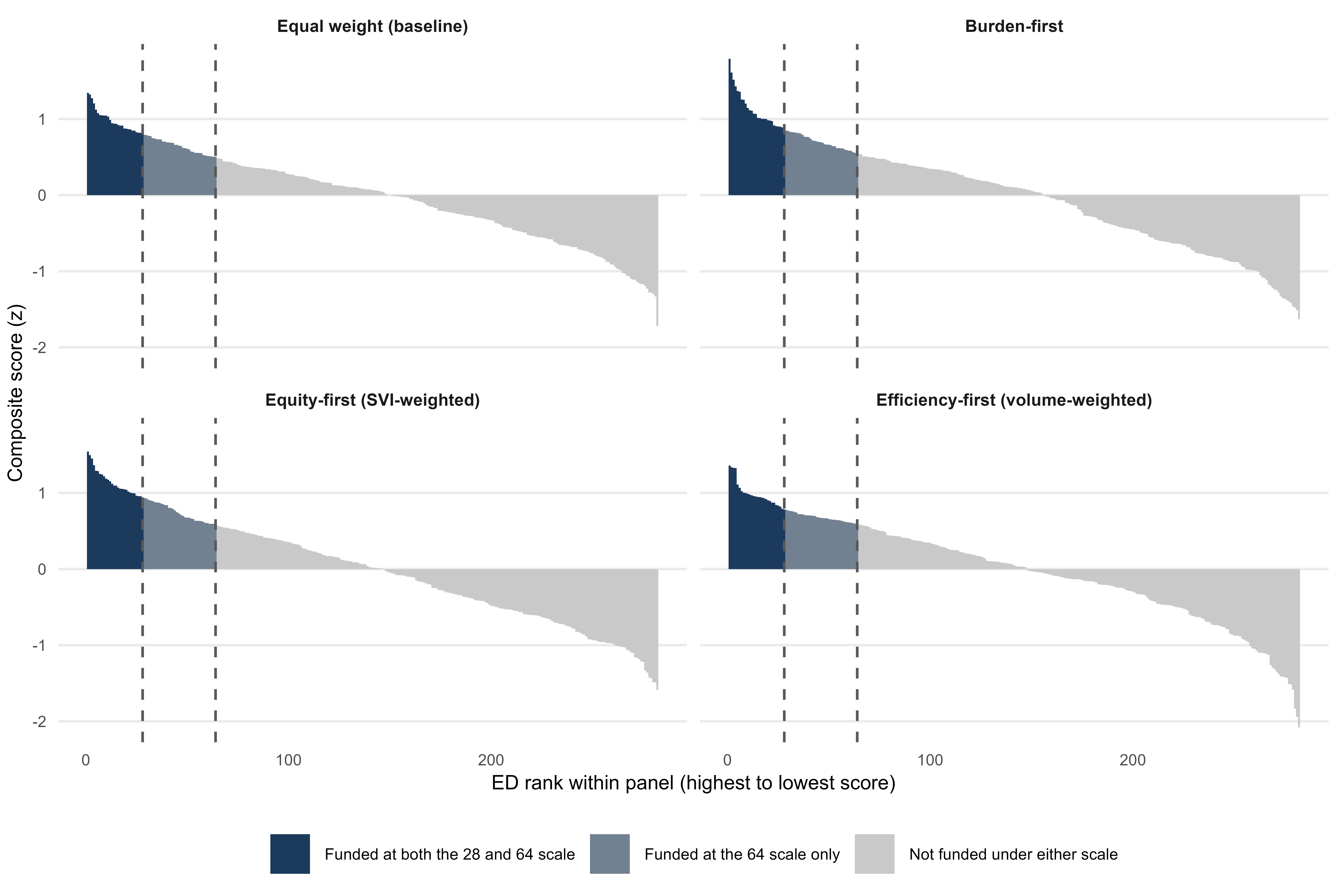

**Legend**: Emergency departments ranked by composite score under equal, burden-first, equity-first, and efficiency-first weightings. Burden represents county syndemic z-score; SVI represents MSSA-level vulnerability; Visits represents log visit volume. Proportions reflect burden, vulnerability, and volume, respectively: equal (1/3, 1/3, 1/3), burden-first (0.6, 0.2, 0.2), equity-first (0.2, 0.6, 0.2), and efficiency-first (0.2, 0.2, 0.6). Dashed vertical lines represent funding cutoffs for the 28 and 64 ED award scale cutoffs.

**Abbreviations**: ED (emergency department), MSSA (Medical Service Study Area), SVI (social vulnerability index score)

**Supplemental Figure 5.** Mean county burden by strategy and participation rate, versus random selection and the 2023 awardees

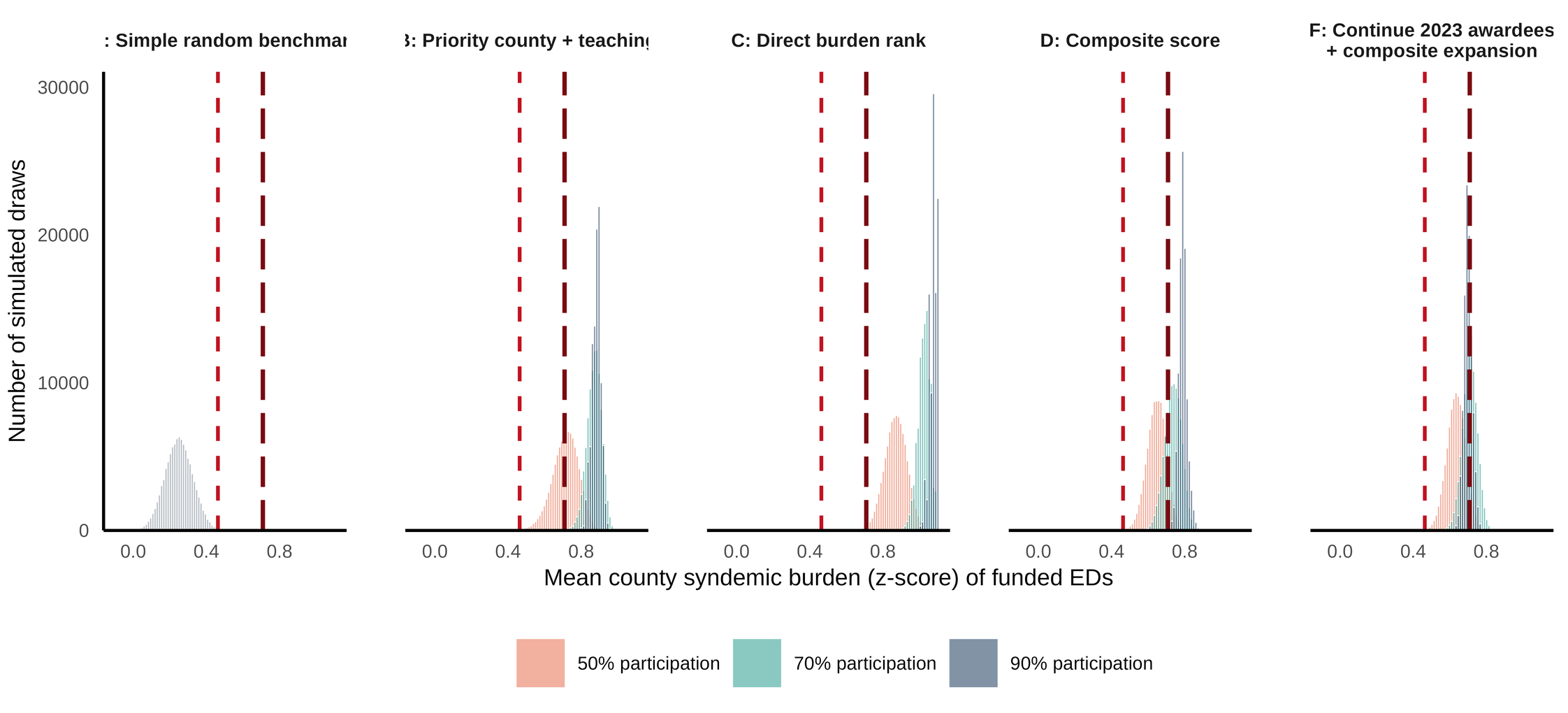

**Legend**: This represents the sensitivity analysis companion to Figure 1, incorporating the 64-emergency department award scale. Distributions reflect mean county syndemic burden for Strategies B–D and F at 50%, 70%, and 90% participation. All at 100,000 draws each. Strategy E is represented by a dashed vertical reference line. Strategy F is represented by the long-dashed dark red vertical line at 70% participation.

**Abbreviations**: ED (emergency department)

**Supplemental Figure 6.** Re-weighting the composite trades disease burden against geographic reach (64-emergency department scale, 70% participation)

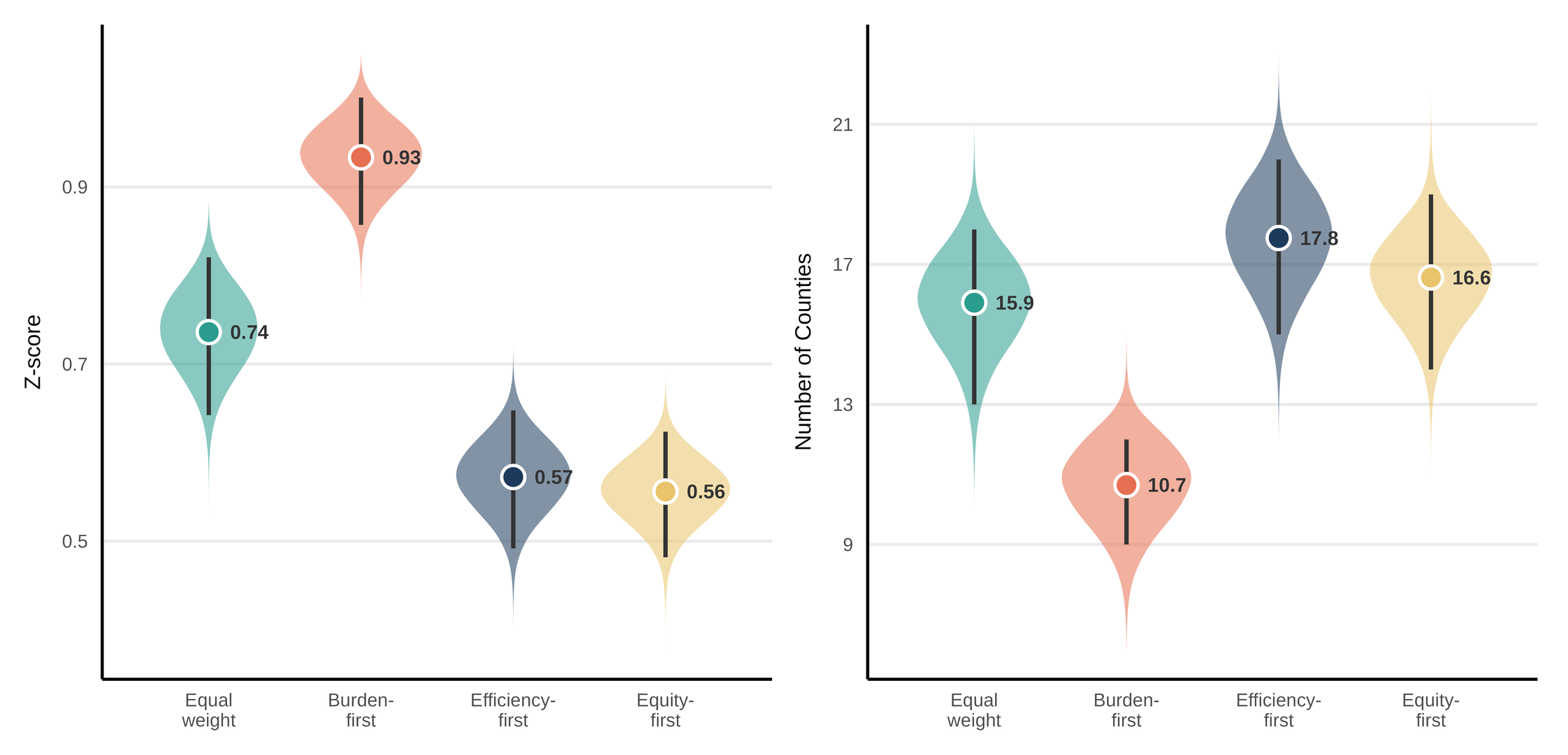

**Legend**: This represents the sensitivity analysis companion to Figure 2, incorporating the 64-emergency department award scale. Mean county burden (left; z-score) and counties reached (right; of 35) under four weightings of Strategy D’s composite at 70% participation. Violins show the full distribution across 100,000 draws. The point marks the mean with corresponding vertical bars representing the 95% simulation interval. Composite weightings of burden, vulnerability, and volume: equal (1/3, 1/3, 1/3), burden-first (0.6, 0.2, 0.2), equity-first (0.2, 0.6, 0.2), and efficiency-first (0.2, 0.2, 0.6).

**Abbreviations**: ED (emergency department)
