## Supplemental Tables for "A Modeling Analysis of Strategies for Allocating Emergency Department HIV and Syphilis Screening Funds in California"

**Supplemental Table 1.** Performance of simulated strategies against actual 2023 awardees, under different participation rates and award scales

| **Award scale** | **Measure** | **B. Priority county + teaching** | | | **C. Direct burden rank** | | | **D. Composite** | | | **F. Continue 2023 awardees + composite expansion** | | |
| --- | --- | --- | --- | --- | --- | --- | --- | --- | --- | --- | --- | --- | --- |
|  |  | **50%** | **70%** | **90%** | **50%** | **70%** | **90%** | **50%** | **70%** | **90%** | **50%** | **70%** | **90%** |
| 28 | Mean county burden (z-score) | 0.89  (0.71-1.06) | 0.89  (0.73-1.05) | 0.78  (0.62-0.97) | 1.13  (0.99-1.27) | 1.24  (1.10-1.36) | 1.34  (1.24-1.40) | 0.82  (0.70-0.94) | 0.85  (0.75-0.93) | 0.91  (0.84-0.96) | 0.69  (0.55-0.83) | 0.63  (0.52-0.77) | 0.56  (0.49-0.64) |
|  | Counties reached (of 35) | 7  (5-8) | 7  (6-8) | 8  (7-8) | 4  (4-4) | 4  (4-4) | 4  (4-4) | 9  (7-12) | 8  (6-10) | 8  (7-8) | 11  (8-14) | 12  (9-14) | 13  (11-14) |
|  | Homeless-patient visits reached (%) | 4.1  (2.9-5.2) | 4.8  (3.6-5.8) | 5.1  (4.5-5.8) | 3.7  (2.6-4.8) | 4.4  (3.2-5.5) | 5.0  (4.3-5.6) | 3.5  (2.6-4.4) | 3.9  (3.0-4.7) | 4.3  (3.7-4.7) | 4.1  (3.0-5.2) | 4.7  (3.8-5.5) | 5.4  (4.5-5.9) |
|  | Uninsured visits reached (%) | 5.1  (4.4-5.8) | 5.1  (4.5-5.7) | 5.4  (5.0-5.8) | 5.0  (4.3-5.7) | 4.9  (4.4-5.5) | 5.3  (4.8-5.6) | 4.9  (4.2-5.7) | 5.1  (4.5-5.8) | 5.0  (4.6-5.4) | 5.2  (4.4-6.0) | 5.2  (4.5-5.9) | 5.6  (5.1-6.1) |
|  | Black-patient visits reached (%) | 14.3  (11.5-16.9) | 15.0  (12.8-16.9) | 14.1  (13.0-15.4) | 13.0  (10.0-15.9) | 13.4  (11.0-15.6) | 11.6  (10.7-14.0) | 12.1  (9.9-14.3) | 11.8  (10.1-13.4) | 12.0  (10.7-12.6) | 12.9  (10.5-15.2) | 13.9  (11.6-15.7) | 15.0  (13.3-16.1) |
|  | Hispanic-patient visits reached (%) | 47.7  (42.6-52.8) | 45.8  (41.5-50.4) | 44.0  (41.3-46.1) | 51.6  (46.3-56.5) | 51.1  (47.0-55.2) | 50.4  (47.2-52.6) | 55.3  (50.7-59.6) | 57.3  (53.4-60.7) | 59.1  (57.3-61.3) | 54.7  (49.9-59.3) | 55.1  (51.6-59.0) | 53.8  (51.4-56.6) |
|  | ED visit volume reached (millions) | 2.23  (1.90-2.51) | 2.31  (2.08-2.55) | 2.37  (2.12-2.52) | 1.81  (1.57-2.05) | 1.75  (1.50-2.00) | 1.54  (1.42-1.68) | 2.21  (1.99-2.43) | 2.23  (2.06-2.40) | 2.21  (2.08-2.30) | 2.03  (1.78-2.29) | 1.99  (1.78-2.19) | 1.89  (1.75-2.10) |
| 64 | Mean county burden (z-score) | 0.71  (0.56-0.84) | 0.87  (0.79-0.94) | 0.88  (0.83-0.93) | 0.87  (0.75-0.98) | 1.02  (0.95-1.08) | 1.08  (1.03-1.10) | 0.65  (0.54-0.75) | 0.74  (0.64-0.82) | 0.79  (0.75-0.84) | 0.63  (0.53-0.73) | 0.71  (0.63-0.78) | 0.70  (0.66-0.75) |
|  | Counties reached (of 35) | 7  (6-8) | 7  (6-8) | 8  (7-8) | 11  (8-13) | 5  (4-7) | 4  (4-4) | 17  (14-20) | 16  (13-18) | 13  (12-13) | 18  (15-21) | 16  (14-19) | 15  (14-17) |
|  | Homeless-patient visits reached (%) | 3.5  (2.8-4.3) | 3.8  (3.2-4.4) | 3.9  (3.5-4.1) | 3.4  (2.7-4.0) | 3.5  (2.9-4.0) | 3.5  (3.2-3.8) | 3.2  (2.6-3.9) | 3.2  (2.8-3.6) | 3.3  (3.0-3.5) | 3.3  (2.7-4.0) | 3.5  (3.0-3.9) | 3.7  (3.4-4.0) |
|  | Uninsured visits reached (%) | 5.0  (4.4-5.5) | 5.3  (4.9-5.7) | 5.2  (5.0-5.5) | 5.0  (4.6-5.5) | 5.2  (4.8-5.6) | 5.2  (5.0-5.4) | 5.0  (4.6-5.5) | 5.0  (4.6-5.4) | 4.9  (4.7-5.2) | 5.1  (4.6-5.6) | 5.0  (4.7-5.4) | 5.0  (4.7-5.2) |
|  | Black-patient visits reached (%) | 12.8  (10.9-14.8) | 13.8  (12.2-15.3) | 14.1  (13.2-14.9) | 12.4  (10.4-14.2) | 12.6  (10.9-14.1) | 13.1  (12.0-13.9) | 11.6  (9.9-13.3) | 12.0  (10.6-13.3) | 12.5  (11.6-13.1) | 11.9  (10.2-13.7) | 12.7  (11.2-14.1) | 12.7  (11.8-13.3) |
|  | Hispanic-patient visits reached (%) | 47.3  (43.5-51.0) | 47.6  (44.5-50.5) | 47.6  (46.0-49.1) | 47.5  (43.4-51.7) | 50.7  (47.5-53.4) | 50.9  (49.2-52.3) | 51.6  (47.8-55.1) | 52.9  (50.4-55.5) | 53.8  (52.2-55.2) | 51.3  (47.6-54.7) | 52.1  (49.5-54.7) | 53.9  (52.4-55.2) |
|  | ED visit volume reached (millions) | 3.81  (3.40-4.22) | 3.99  (3.58-4.44) | 4.68  (4.39-4.89) | 3.49  (3.10-3.90) | 3.37  (3.03-3.74) | 3.91  (3.68-4.08) | 4.27  (3.90-4.64) | 4.63  (4.33-4.91) | 4.93  (4.71-5.08) | 4.21  (3.84-4.59) | 4.56  (4.22-4.86) | 4.71  (4.55-4.88) |

**Legend**: Strategies B, C, D, and F at 50% (conservative), 70% (primary), and 90% (optimistic) participation at the 28- and 64-emergency department award scales. Values are presented as means with 95% simulation interval [SI] across 100,000 draws. ED visit volume is shown in millions of annual patient visits.

**Abbreviations**: ED (emergency department). SI (simulation interval)

**Supplemental Table 2.** Performance of simulated strategies at the 64-emergency department award scale

| **Measure** | **A. Simple random benchmark** | **B. Priority county + teaching** | **C. Direct burden rank** | **D. Composite** | **F. Continue 2023 awardees + composite expansion** |
| --- | --- | --- | --- | --- | --- |
| Counties reached (of 35) | 24 (20-28) | 7 (6-8) | 5 (4-7) | 16 (13-18) | 16 (14-19) |
| Mean county burden (z-score) | 0.25 (0.10-0.40) | 0.87 (0.79-0.94) | 1.02 (0.95-1.08) | 0.74 (0.64-0.82) | 0.71 (0.63-0.78) |
| Mean social vulnerability (MSSA SVI, 0-1) | 0.51 (0.46-0.55) | 0.52 (0.50-0.55) | 0.54 (0.52-0.58) | 0.66 (0.64-0.68) | 0.64 (0.62-0.67) |
| Homeless-patient visits reached, n (%) | 99,481 (2.9%)  [2.2-3.7%] | 153,675 (3.8%)  [3.2-4.4%] | 118,075 (3.5%)  [2.9-4.0%] | 148,629 (3.2%)  [2.8-3.6%] | 159,584 (3.5%)  [3.0-3.9%] |
| Uninsured visits reached, n (%) | 165,262 (4.9%)  [4.3-5.5%] | 210,584 (5.3%)  [4.9-5.7%] | 175,509 (5.2%)  [4.8-5.6%] | 230,438 (5.0%)  [4.6-5.4%] | 229,430 (5.0%)  [4.7-5.4%] |
| Black-patient visits reached, n (%) | 342,335 (10.0%)  [7.9-12.3%] | 551,859 (13.8%)  [12.2-15.3%] | 426,670 (12.6%)  [10.9-14.1%] | 556,629 (12.0%)  [10.6-13.3%] | 580,581 (12.7%)  [11.2-14.1%] |
| Hispanic-patient visits reached, n (%) | 1,517,851 (44.6%)  [39.8-49.3%] | 1,902,872 (47.6%)  [44.5-50.5%] | 1,709,385 (50.7%)  [47.5-53.4%] | 2,447,245 (52.9%)  [50.4-55.5%] | 2,374,690 (52.1%)  [49.5-54.7%] |
| ED visit volume reached, n (%) | 3,404,700 (22.7%)  [19.7-25.8%] | 3,993,982 (26.6%)  [23.9-29.6%] | 3,372,520 (22.5%)  [20.2-24.9%] | 4,625,879 (30.8%)  [28.8-32.7%] | 4,555,785 (30.4%)  [28.1-32.4%] |

**Legend**: This represents the sensitivity analysis companion to Table 1 at the 64-emergency department award scale, adding Strategy F. Ranked Strategies B-D and F are modeled at 70% participation. Values are presented as means with 95% simulation interval [SI] across 100,000 draws.

**Abbreviations**: ED (emergency department), MSSA (Medical Service Study Area), SVI (social vulnerability index)
